# A polygenic risk score identifies undiagnosed cases of diabetes

**DOI:** 10.1101/2023.10.31.23297847

**Authors:** Chris German, James Ashenhurst, Wei Wang, 23andMe Research Team, Julie M. Granka, Bertram L. Koelsch, Noura S. Abul-Husn, Stella Aslibekyan, Adam Auton, Joyce Tung, Suyash S. Shringarpure, Michael V. Holmes

**Affiliations:** 23andMe, Sunnyvale, CA, USA

## Abstract

**Importance:** Twenty-three percent of 37.3M adults in the USA with diabetes are estimated to be undiagnosed, leading to potentially avoidable sequelae and morbidity.

**Objective:** To explore the utility of a polygenic risk score (PRS) at identifying individuals with undiagnosed diabetes and prediabetes.

**Design, Setting and Participants:** Individuals without doctor-diagnosed diabetes at study baseline in the UK Biobank (UKB) with HbA1c and BMI measurements. Participants were restricted to white individuals to use an ancestry-appropriate PRS. Undiagnosed diabetes and prediabetes were defined using HbA1c (≥6.5% and ≥5.7 - <6.5%, respectively).

**Exposures:** A diabetes PRS comprising 13,863 SNPs derived from the 23andMe Research Cohort, and measured BMI among UKB participants.

**Results:** Of 412,439 individuals self-reporting an absence of diagnosed diabetes and who had BMI and HbA1c measurements at baseline, 2,934 (0.7%) had undiagnosed diabetes, representing 11.9% of all (diagnosed and undiagnosed) diabetes. Nearly half (1,362, 46%) of undiagnosed diabetes cases were among individuals in the top 25% of the PRS distribution. Overweight individuals (BMI ≥25 - <30 kg/m^2^) who were in the top 12.5% of the PRS distribution had a similar frequency of undiagnosed diabetes (0.8-1.6% frequency) as individuals with obesity (BMI ≥30kg/m^2^) in the lowest 12.5% of the PRS distribution (0.7-1.7% frequency). Combining overweight and obesity with the PRS identified nearly all cases of undiagnosed diabetes: individuals with a BMI ≥25 kg/m^2^ (66% of the study population) or those in the top 54-69% of the PRS identified 98-99% of undiagnosed cases. Of the 199 undiagnosed diabetes cases occurring among individuals with a normal BMI (<25kg/m^2^), two-thirds were among individuals in the top 50% of the PRS. Prediabetes was common (14%), with measured BMI and PRS providing additive risk. Among those in the top 12.5% PRS with BMI ≥35kg/m^2^, 6.3% developed incident diabetes over 4 years follow-up, as compared to 0% among the bottom 12.5% PRS with BMI<25kg/m^2^.

**Conclusions:** A diabetes PRS is informative at identifying undiagnosed cases. PRS may have broader utility in detecting individuals with asymptomatic disease.

**Key Points:** *Question:* Does a polygenic risk score (PRS) have utility in identifying individuals with undiagnosed type 2 diabetes (T2D)?

*Findings:* In this analysis of 412,439 individuals without doctor-diagnosed diabetes, a T2D PRS performed additively to body mass index (BMI) at identifying individuals with undiagnosed diabetes. Selecting individuals on the basis of overweight/obesity or a T2D PRS identified almost all cases of undiagnosed diabetes. The majority of undiagnosed diabetes cases among individuals with normal weight occurred among those at elevated polygenic risk.

*Meaning:* A T2D PRS identifies cases of undiagnosed diabetes among individuals with and without overweight or obesity.

## Introduction

Diabetes is among the top 10 leading causes of death and disability worldwide^1^. Individuals with diabetes are at twice the risk of dying from any cause^2^, and are at a nearly three-fold higher risk of death from cardiovascular disease ^3^.

The incidence and prevalence of diabetes has doubled since the 1990s^4^, mostly driven by increases in population weight^5^. In 2019, there were 37.3M adults with diabetes in the United States, representing 11.3% of the adult population^6^. Notably, it is estimated by the American Diabetes Association (ADA) that almost 1 in 4 adults with diabetes are not aware of or did not report having diabetes, i.e. that their diabetes is undiagnosed^6^, with more recent estimates putting this closer to one in ten^7^. Early exposure to dysglycemia^8^ and time to target HbA1c^9^ are associated with elevated risk of diabetes-related complications and mortality^10^, making the early detection of undiagnosed diabetes a potentially important public health goal. The ADA^11^ recommends screening for prediabetes and diabetes in all individuals over 35 years, and the US Preventive Services Task Force (USPSTF) additionally recommends screening for individuals overweight or with obesity^12^. Such guidelines differ between countries - for example, the UK National Screening Committee (UK NSC) does not currently recommend screening for diabetes^13^.

Polygenic risk scores (PRS) aggregate the effects of thousands of genetic variants associated with a trait such as a health outcome and can be used to identify individuals at risk of disease^14^. One unexplored clinical application is the use of PRS in identifying individuals with undiagnosed disease. In this study, we explored the clinical utility of aT2D PRS to identify individuals with undiagnosed diabetes.

## Methods

### Study Cohort

We used data from the UK Biobank (UKB), a prospective cohort of ∼0.5M adults from the UK^15^, under Application Number 95801. Participant exclusions (i.e. removing individuals who no longer provided consent) were conducted as per recommendations. Details on genotyping quality control, phasing, and imputation in UKB have been described previously^15^.

### Construction of the T2D PRS

We used data from the 23andMe, Inc. Research Cohort^16^ to construct a T2D associated PRS. Individuals included were research participants of 23andMe, Inc., a direct-to-consumer genetics company, who were genotyped as part of the 23andMe Personal Genome Service. Participants provided informed consent and volunteered to participate in the research online, under a protocol approved by the external AAHRPP-accredited IRB, Ethical & Independent (E&I) Review Services. As of 2022, E&I Review Services is part of Salus IRB (https://www.versiticlinicaltrials.org/salusirb).

Development of the utilized PRS has been previously described^17^. Briefly, we first conducted GWAS in 5 ancestry groups separately (European, Hispanic/Latino, Sub-Saharan African/African American, East/Southeast Asian, and South Asian) and then conducted a meta-analysis of the GWAS summary statistics. In addition to rs7903146, a variant with strong evidence of association with T2D^18^, SNPs were selected from this meta-analysis. The PRS model was trained on a mixed-ancestry training cohort, including individuals of European, Hispanic/Latino, and Sub-Saharan African/African American descent. To select variant sets, we performed pruning and thresholding with combinations of selection hyperparameters. For example: distance (kb) = [10, 100, 200, 1000, 2000], and GWAS p-value = [1e-2, 1e-4, 1e-6, 1e-8]. We fitted models for each SNP set with age, sex, age^2^, sex * age interaction, and sex * age^2^ interaction, as well as the first ten global principal components (PCs) to account for population substructure. We used Scikit-learn’s LogisticRegression gradient descent algorithms^19^ to determine optimal parameter weights using the liblinear solver and L2 regularization. The final model was composed of a single set of variants selected based on performance in an ancestry-specific European validation cohort. This led to a final PRS model of 13,869 SNPs that, after overlapping with variants in the UK Biobank, consisted of 13,863 SNPs.

### Exclusion criteria

We restricted analyses to individuals in the UKB who had non-missing data on self-reported doctor-diagnosed diabetes, non-missing BMI data, and laboratory values for HbA1c at instance 0 (study baseline). Analyses were additionally restricted to individuals self-reporting white ethnicity at baseline.

### Outcomes

We transformed HbA1c from mmol/mol to % using the formula HbA1c % = (HbA1c (mmol/mol) / 10.929) + 2.15.^20^ We defined undiagnosed diabetes and prediabetes among individuals with an absence of prior diagnosed diabetes. Prior diagnosed diabetes used a combination of self-reported doctor-diagnosed diabetes at instance 0 (baseline) and first occurrence data (data fields 130706 through 130715, see **Table S1**). We defined undiagnosed diabetes as those with an HbA1c ≥ 6.5% at instance 0 (baseline). Prediabetes was defined as individuals with an A1c % of ≥ 5.7% and <6.5%^11^ at instance 0. Separately, we defined those that developed incident diabetes during follow-up (either self-report or by HbA1c ≥ 6.5% at instance 1) among people without diabetes at instance 0 and whose HbA1c was <6.5% at instance 0.

### Statistical analysis

We quantified the number of individuals with undiagnosed diabetes and prediabetes based on HbA1c levels among all individuals who reported ‘no’ to doctor diagnosed diabetes at baseline and did not have a first occurrence diagnosis of diabetes prior to baseline. This was established using the HbA1c cut-points described above, using samples measured at baseline (instance 0) in 412,439 individuals. These provided the number of individuals with undiagnosed diabetes and prediabetes in the cohort. The total number of individuals with diabetes was calculated as the number of individuals either reporting doctor diagnosed diabetes at baseline or with a confirmed first occurrence diagnosis of diabetes with a date prior to baseline, together with the number of individuals with undiagnosed diabetes at baseline.

We then estimated the number of individuals with undiagnosed diabetes and prediabetes among individuals in the top 25% of the T2D PRS distribution, and in each of 8 bins of the T2D PRS distribution accounting for 12.5% of the distribution (i.e. octiles). For each of these groups, we quantified the number of individuals with undiagnosed diabetes or prediabetes divided by the total number of individuals in the group.

We evaluated the performance of the T2D PRS with body mass index (BMI) using measured BMI (derived from weight and height) rather than BMI derived from body impedance. First, we quantified the number of individuals with undiagnosed diabetes across the distribution of BMI and among those overweight or with obesity (BMI ≥25kg/m^2^). Second, we restricted our analysis to those overweight or with obesity (BMI ≥25kg/m^2^) and explored the additive effect of identifying individuals with undiagnosed diabetes by also including individuals based on T2D PRS thresholding. Third, we compared the performance of the T2D PRS and BMI at identifying individuals with undiagnosed diabetes using the same denominator (number of individuals in whom HbA1c would be used to establish undiagnosed diabetes). 65.6% of UKB participants had a BMI ≥25kg/m^2^; therefore, we selected a PRS threshold of the top 65.6% for this analysis. We then tested for the presence of undiagnosed diabetes in each group, in which the numerators were equally sized (i.e., 65.6% of the total population).

Similar analyses were conducted with prediabetes and incident diabetes (defined above) as the outcome. We conducted sensitivity analyses stratified by age (<50 years, ≥50 to <60 years, ≥60 years) and self-reported sex.

Analyses were performed using R version 4.1.2.

## Results

### Study cohort and diabetes (diagnosed, undiagnosed and prediabetes)

Among 434,110 self-reported white individuals in the UK Biobank who had BMI and HbA1c measurements at baseline and had an informative (Yes/No) answer to doctor-diagnosed diabetes, 20,900 (4.8%) self-reported presence of doctor-diagnosed diabetes at baseline. Checking first occurrence data for all types of diabetes prior to initial assessment visit at instance 0 resulted in 771 additional cases, leading to a total 21,671 (5.0%) cases of prevalent diabetes.

Of 412,439 individuals without doctor-diagnosed diabetes at baseline, the average HbA1c was 5.4% (SD: 0.4) and BMI was 27.2 kg/m^2^ (SD: 4.6). The average age of included individuals was 57 years, and 55% of study participants were female. A total of 2,934 individuals (0.7%) had an HbA1c ≥6.5%, indicative of undiagnosed diabetes, and 57,317 (13.9%) had an HbA1c ≥5.7% and <6.5% indicative of prediabetes (**Figures 1A, S8**). This corresponds to an estimated 11.9% of all individuals with diabetes being undiagnosed calculated as the proportion of individuals with undiagnosed diabetes (2,934) divided by the total number of individuals with diabetes (the sum of 2,934 undiagnosed cases and 21,671 prevalent diabetes cases). **Table 1** shows characteristics of individuals by whether they had undiagnosed diabetes or prediabetes vs. individuals free of either. Fewer females than males (40% vs 55%) had undiagnosed diabetes, and undiagnosed diabetes cases were more likely to have ever smoked (68% vs 60%), had a higher BMI (32kg/m^2^ vs 27kg/m^2^), and higher systolic (147mmHg vs 137mmHg) and diastolic (87mmHg vs 82mmgHg) blood pressures as compared to individuals free of undiagnosed diabetes or prediabetes. **Figure S7** shows the distribution of the T2D PRS values based on diabetes status. Individuals with diagnosed and undiagnosed diabetes followed a more similar distribution than individuals free of diabetes or prediabetes.

**Figure 1:**
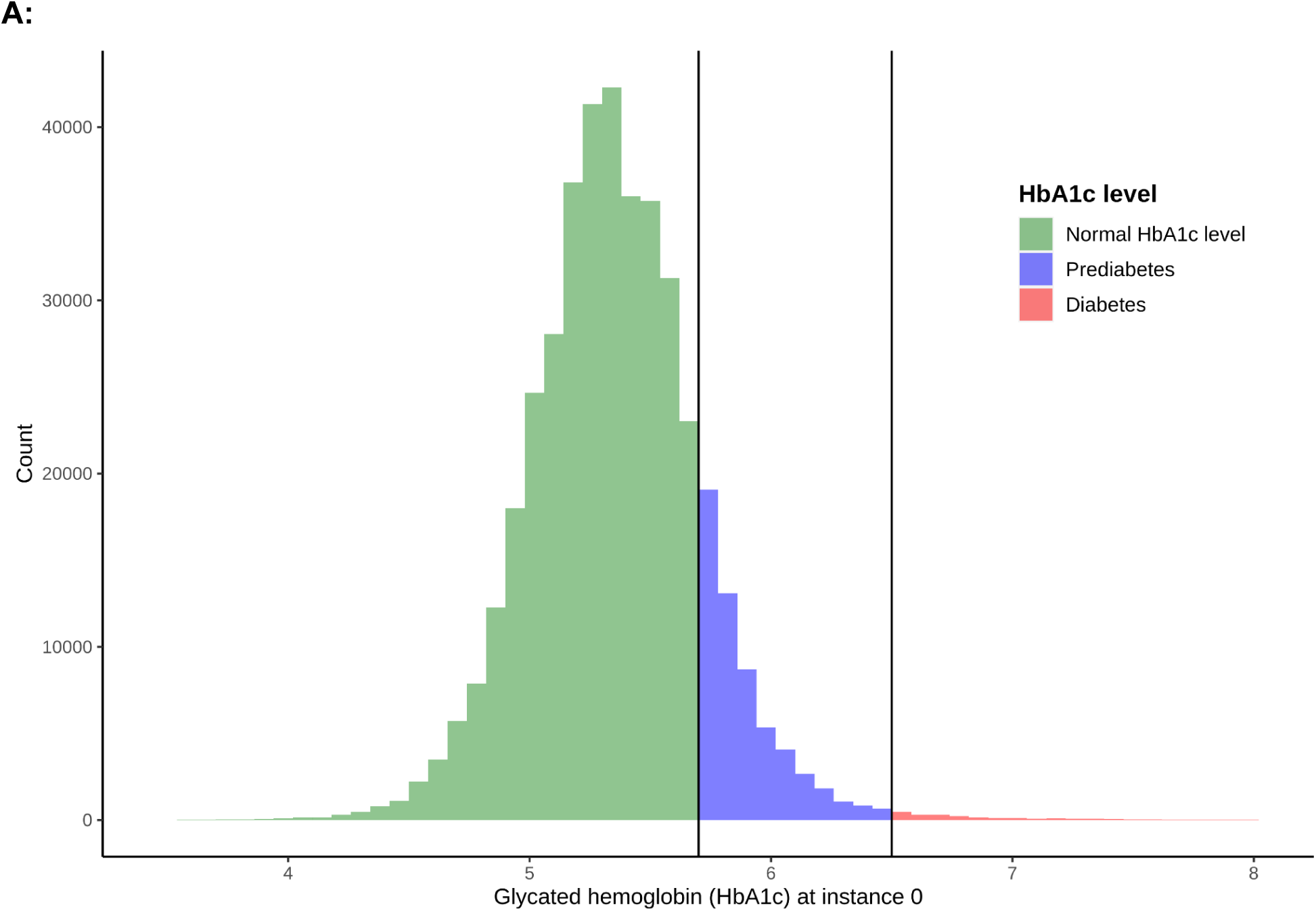
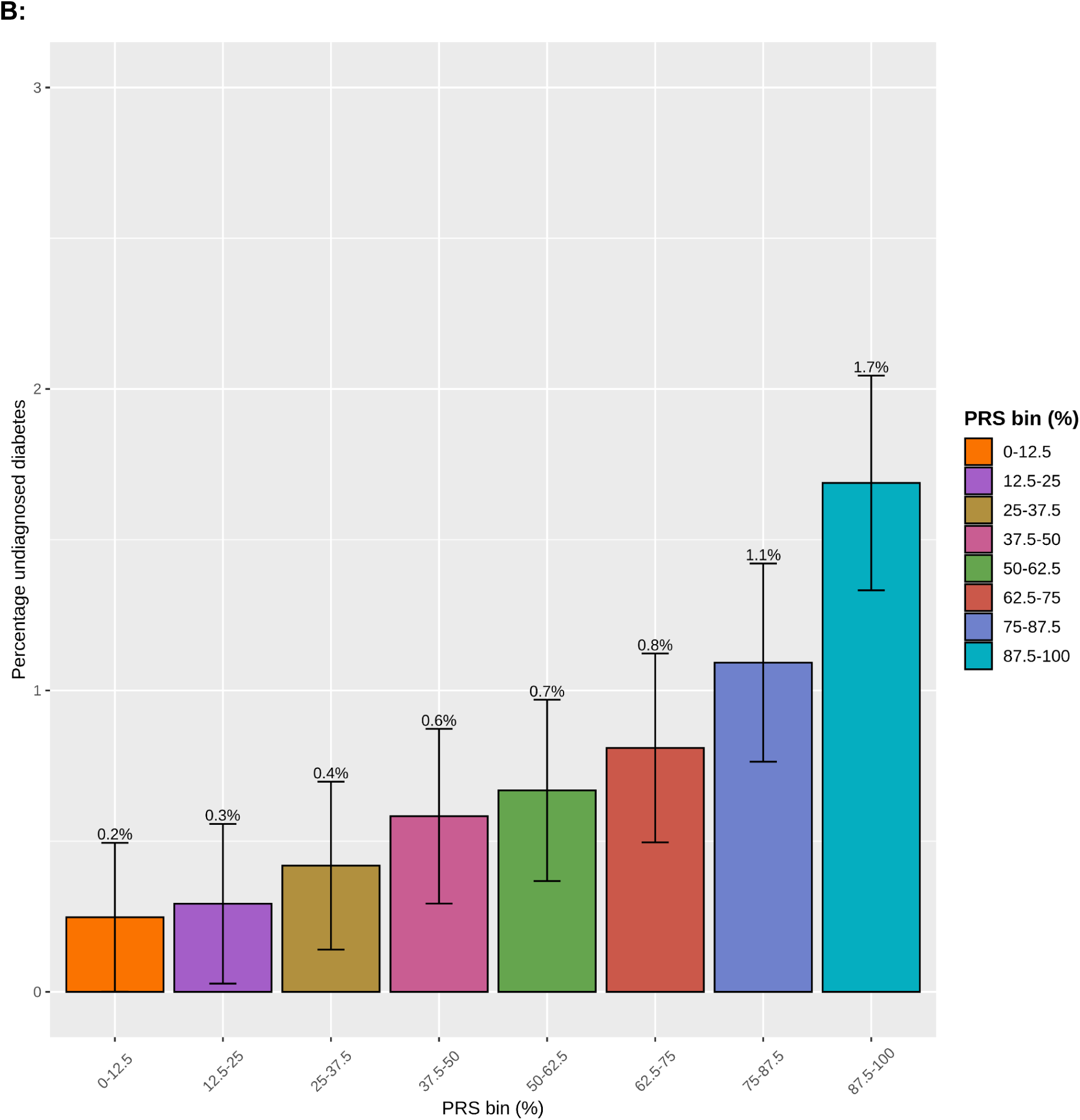
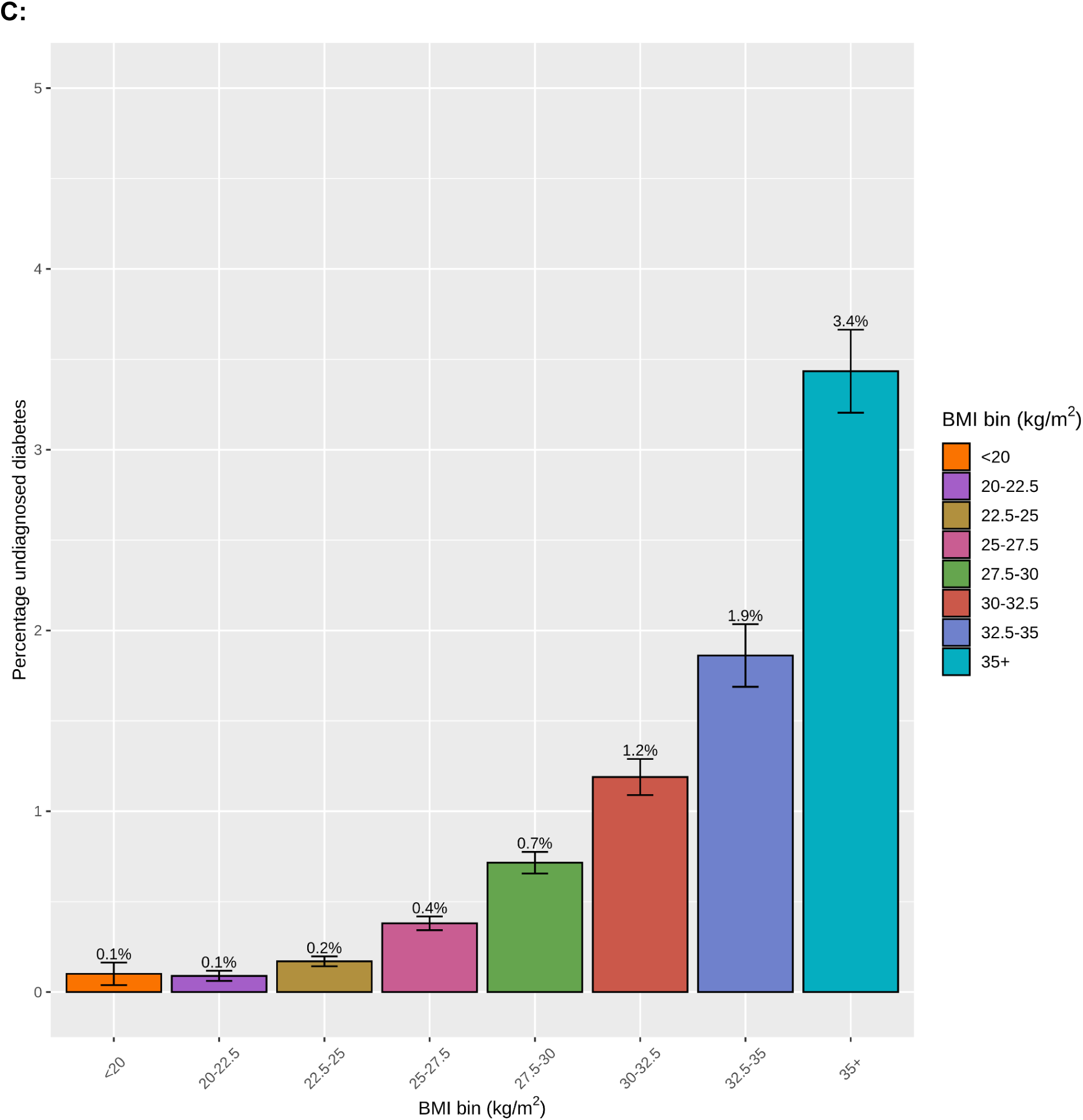
Undiagnosed diabetes and prediabetes by HbA1c values (A), and frequency of undiagnosed diabetes by strata of (B) T2D PRS and (C) body mass index. HbA1c was transformed from mmol/mol to % using (HbA1C (mmol/mol) / 10.929) + 2.15^14^. Histogram of HbA1c is right-truncated at 8%. T2D PRS in B is derived from 23andMe. Error bars in B and C relate to 95% confidence intervals. Bins in B and C are left-inclusive - e.g. BMI 20-22.5 includes all individuals with a BMI of 20kg/m^2^ to less than 22.5kg/m^2^.

**Table 1.** Characteristics of individuals with undiagnosed diabetes and prediabetes in the UK Biobank. Restricted to individuals without doctor-diagnosed diabetes. Given the T2D PRS derived from 23andMe was trained on individuals of European genetic ancestry, we additionally restricted to individuals self-reporting white ethnicity.

|  | **Undiagnosed diabetes** | **Prediabetes** | **Individuals free of diabetes or prediabetes** |
| --- | --- | --- | --- |
| Number | 2,934 | 57,317 | 352,188 |
| Age, mean (SD) | 60 (7.1) | 61 (6.7) | 56 (8.1) |
| Sex, % Female | 40% | 55% | 55% |
| Ever smoked, % Yes | 68% | 65% | 60% |
| BMI, kg/m<sup>2</sup>, mean (SD) | 32 (5.8) | 29 (5.2) | 27 (4.4) |
| HbA1c, %, mean (SD) | 7.5 (1.7) | 5.9 (0.17) | 5.3 (0.27) |
| SBP, mmHg, mean (SD) | 147 (19) | 142 (19) | 137 (19) |
| DBP, mmHg, mean (SD) | 87 (11) | 83 (10) | 82 (10) |
| Years education, mean (SD) | 14 (5.3) | 14 (5.3) | 15 (5) |

Footnote: years of education was calculated using a previously-published formula<sup>33</sup>. BMI, HbA1c and blood pressure values are from instance 0 (baseline). BMI: body mass index; DBP: diastolic blood pressure; HbA1c: Hemoglobin A1c; SBP: systolic blood pressure.

### T2D PRS, overweight and obesity, and undiagnosed diabetes

Grouping individuals by their T2D PRS identified a dose-response log-linear relationship between polygenic risk and frequency of undiagnosed diabetes (**Figure 1B**). While the population average frequency of undiagnosed diabetes was 0.7%, individuals in the lowest PRS octile (bottom 12.5%) had a 0.2% risk of undiagnosed diabetes as compared to individuals in the highest PRS octile (top 12.5%) who had a frequency of 1.7%, conveying a relative risk (RR) of 6.8 (95%CI: 5.7, 8.2) of undiagnosed diabetes across the extremes of the PRS distribution. Almost half (1,362, 46%) of undiagnosed diabetes cases were contained with the top two octiles (top 25%) of the T2D PRS.

As with the T2D PRS, a dose-response log-linear relationship was identified between measured BMI and risk of undiagnosed diabetes (**Figure 1C**): those with a BMI<20 kg/m^2^ had a 0.1% risk of undiagnosed diabetes whereas those with BMI ≥35kg/m^2^ had a 3.4% frequency. This corresponded to a RR of 34.1 (95%CI: 18.6, 62.7) comparing individuals with BMI ≥35kg/m^2^ vs. BMI <20kg/m^2^. Among 66% (n=270,493) of the population overweight or with obesity (BMI ≥25kg/m^2^), there were 2,735 undiagnosed diabetes cases, representing 93% of all undiagnosed diabetes cases.

We identified separate, additive effects of measured BMI and the T2D PRS on the risk of undiagnosed diabetes, with the underlying BMI amplifying the effect of the T2D PRS (**Figures S3-S4**). As stated above, on average the difference in frequency of undiagnosed diabetes between the top and bottom 12.5% of the PRS distribution was 1.5% (**Figure 1B**). The same comparison of top and bottom PRS octiles was associated with a 0.3% difference in frequency of undiagnosed diabetes among those with a BMI<20 kg/m^2^, 0.7% among those with BMI ≥25-<27.5 kg/m^2^, 1.6% among those with BMI ≥30-<32.5 kg/m^2^, and 4.1% among those with BMI ≥35 kg/m^2^ (**Figure S3**).

Among those with the highest BMI in the UK Biobank (BMI ≥35kg/m^2^), the frequency of undiagnosed diabetes was 3.4% (**Figure 1C**). For those in this same BMI category who were also in the lowest 12.5% of the T2D PRS, the risk was 1.7% (**Figure S3**), which was identical to the risk in the overall population at the highest genetic risk (top 12.5%) who had an average BMI 8kg/m^2^ lower at 27 kg/m^2^. In contrast, those with the highest BMI (≥35kg/m^2^) and at highest polygenic risk of T2D (top 12.5% PRS) had a 5.8% frequency of undiagnosed diabetes, representing a 8.8-fold higher relative risk (95%C: 7.8, 10.1) as compared to the rest of the population, and a 54.9-fold RR compared to those at lowest risk (BMI<20kg/m^2^ and lowest 12.5% T2D PRS).

Figures 2A and **S4** illustrate the separate and additive frequency of undiagnosed diabetes by categories of BMI and T2D PRS. Overweight individuals (BMI 25-30 kg/m2) who were in the top 12.5% of the T2D PRS had a similar frequency of undiagnosed diabetes (0.8-1.6% frequency) as individuals with obesity (BMI ≥30kg/m^2^) in the lowest 12.5% of the T2D PRS (0.7-1.7% frequency).

**Figure 2.**
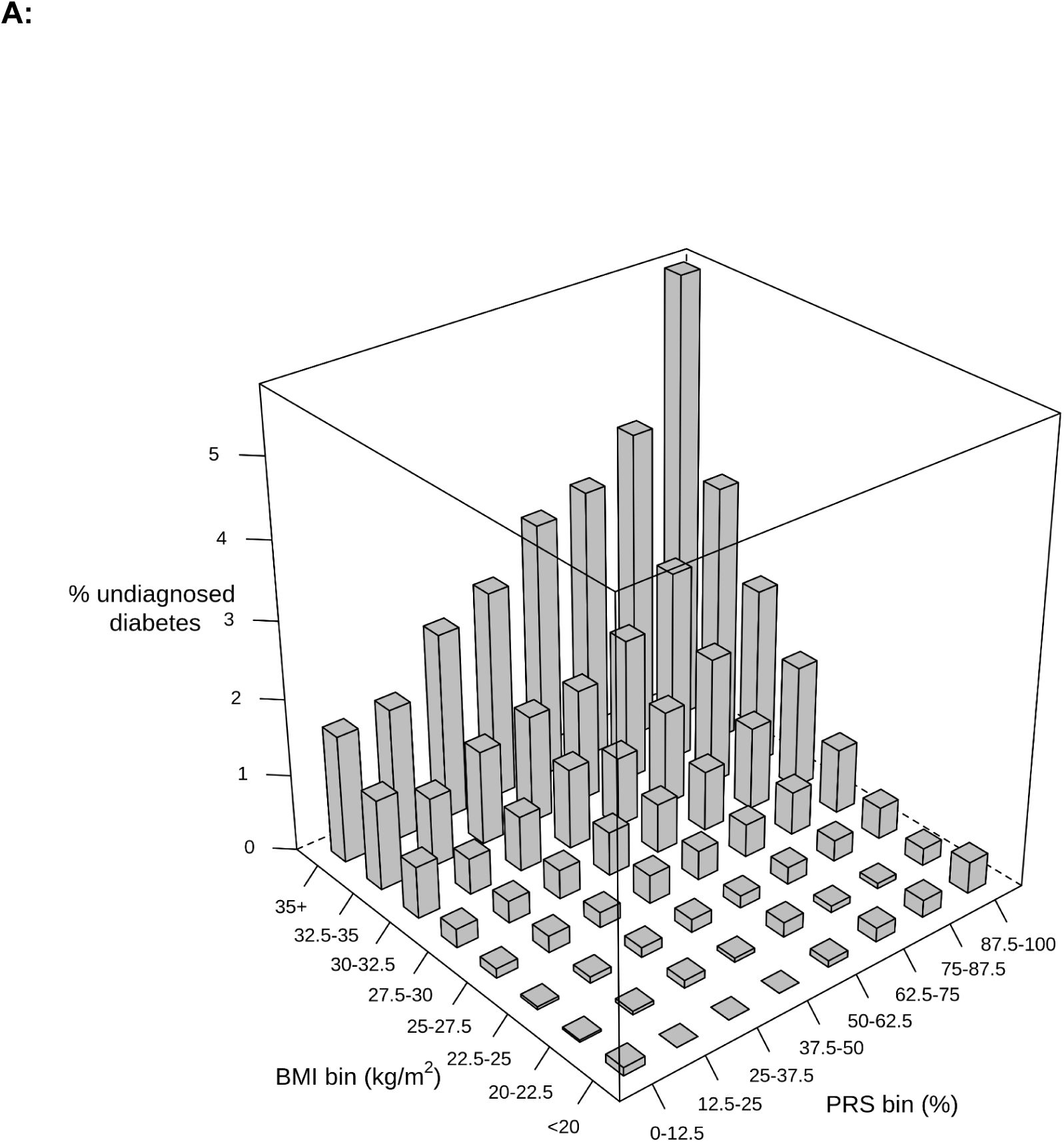
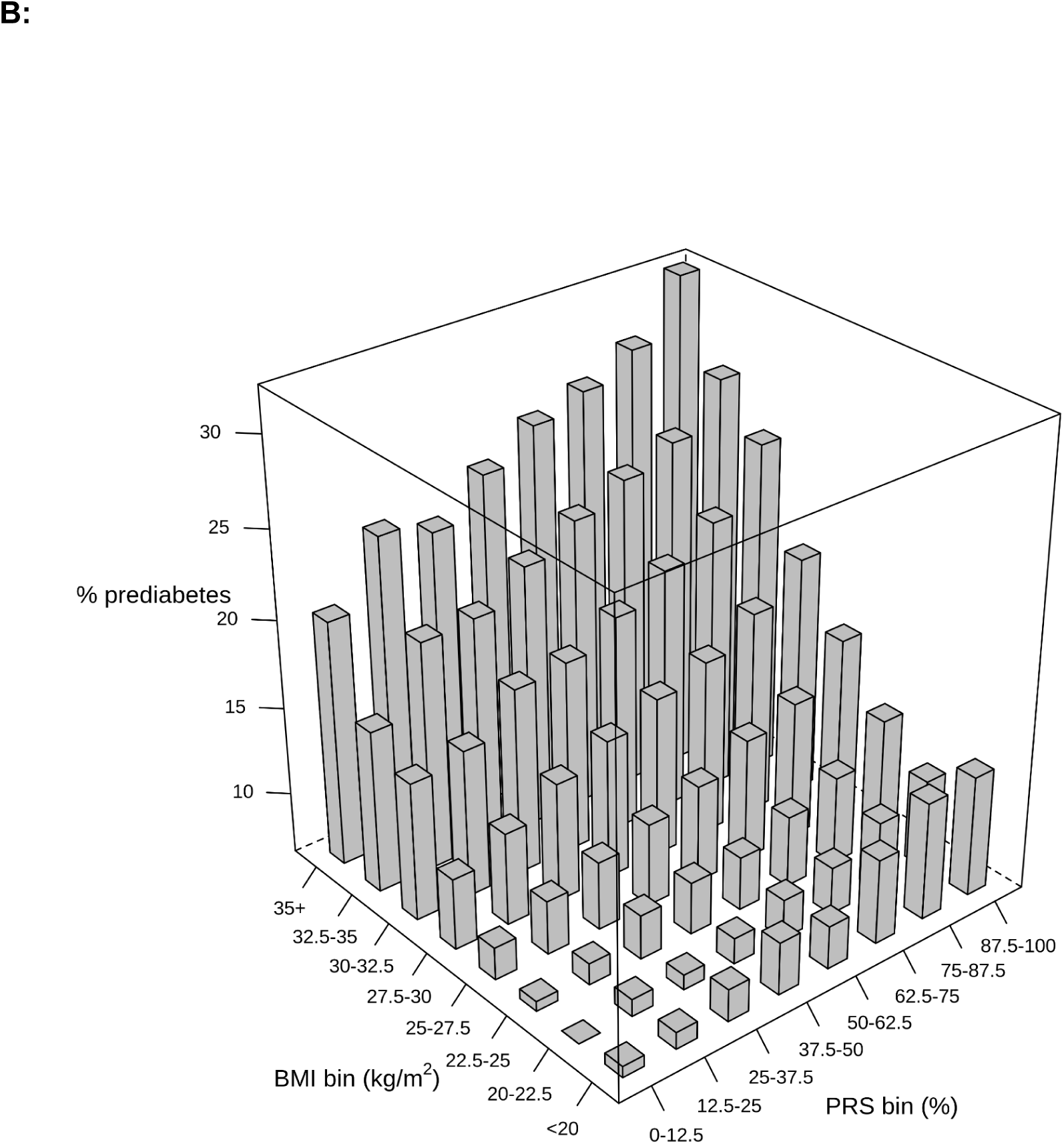
Frequency of (A) undiagnosed diabetes and (B) prediabetes by strata of T2D PRS and body mass index. Individuals were grouped by their T2D PRS into bins of 12.5%. BMI values are in kg/m^2^. The bins are left-inclusive - e.g. BMI 20-22.5 includes all individuals with a BMI of 20kg/m^2^ to less than 22.5kg/m^2^.

Selecting individuals purely on the basis of measured BMI neglected to identify individuals with undiagnosed diabetes arising from the elevated polygenic risk of diabetes. Among individuals with a normal or low BMI (<25kg/m^2^), there were 199 cases of undiagnosed diabetes, corresponding to 6.8% of all undiagnosed diabetes cases. Out of the 199 cases of undiagnosed diabetes among individuals with a normal or low BMI, 77 (38.7%) occurred among individuals in the top 25% of the T2D PRS and 134 (67.3%) among individuals in the top 50% of the T2D PRS.

We explored two complementary approaches to establishing the additive and comparative performance of BMI and the T2D PRS in identifying individuals with undiagnosed diabetes. First, we restricted our focus to individuals who were overweight or obese (BMI ≥25kg/m^2^, roughly 66% of all individuals), which captured 93% of undiagnosed diabetes cases. In an effort to increase the identification of individuals with undiagnosed diabetes, we explored the effects of also including individuals based on thresholding of the T2D PRS (**Table S2**). For example, in addition to those with BMI ≥25kg/m^2^, also including individuals in the top 5% of the T2D PRS identified 94% of cases of undiagnosed diabetes. Broadening this to BMI ≥25kg/m^2^ and the top 26% T2D PRS identified 96% of undiagnosed diabetes, while BMI ≥25kg/m^2^ and the top 54% T2D PRS identified 98% of undiagnosed diabetes. Next, we conducted a more direct comparison by sampling individuals in the T2D PRS by the same cohort proportion who were overweight or obese. Taking BMI ≥25kg/m^2^ comprises 65.6% of the UKB population and so we identified individuals in the top 65.6% of the T2D PRS (**Table S3**). As stated above, using a BMI threshold of ≥25kg/m^2^ detected 93% of undiagnosed diabetes cases. In contrast, among individuals in the top 65.6% of the T2D PRS, 85% of undiagnosed diabetes cases were identified. Collectively, this demonstrates that while measured BMI identifies a slightly larger proportion of cases of undiagnosed diabetes than a comparable thresholding using a T2D PRS, the combination of BMI and T2D PRS can identify the vast majority of undiagnosed diabetes cases.

### Prediabetes and incident diabetes during follow-up

57,317 (13.9%) of individuals had prediabetes at baseline. An additive effect of measured BMI and polygenic risk of T2D was identified on frequency of prediabetes (Figures 2B and **S5**). In individuals with BMI ≥35kg/m^2^ and in the top 12.5% of the T2D PRS distribution, the frequency of prediabetes was nearly 1 in 3 (32.5%). In contrast, this was 7.2% in those with low BMI (<20kg/m^2^) and low T2D PRS (bottom 12.5%).

At follow-up, there were 238 new cases of diabetes (either by self-report or using HbA1c), representing an average incidence of 1.4%. This value was highest among those with both high BMI (≥35kg/m^2^) and high T2D PRS (top 12.5%), with 6.3% of these individuals developing diabetes within 4 years of follow-up (Figure 3). In contrast, no incident cases occurred among those with BMI <25kg/m^2^ who were in the lowest T2D PRS octile.

**Figure 3.**
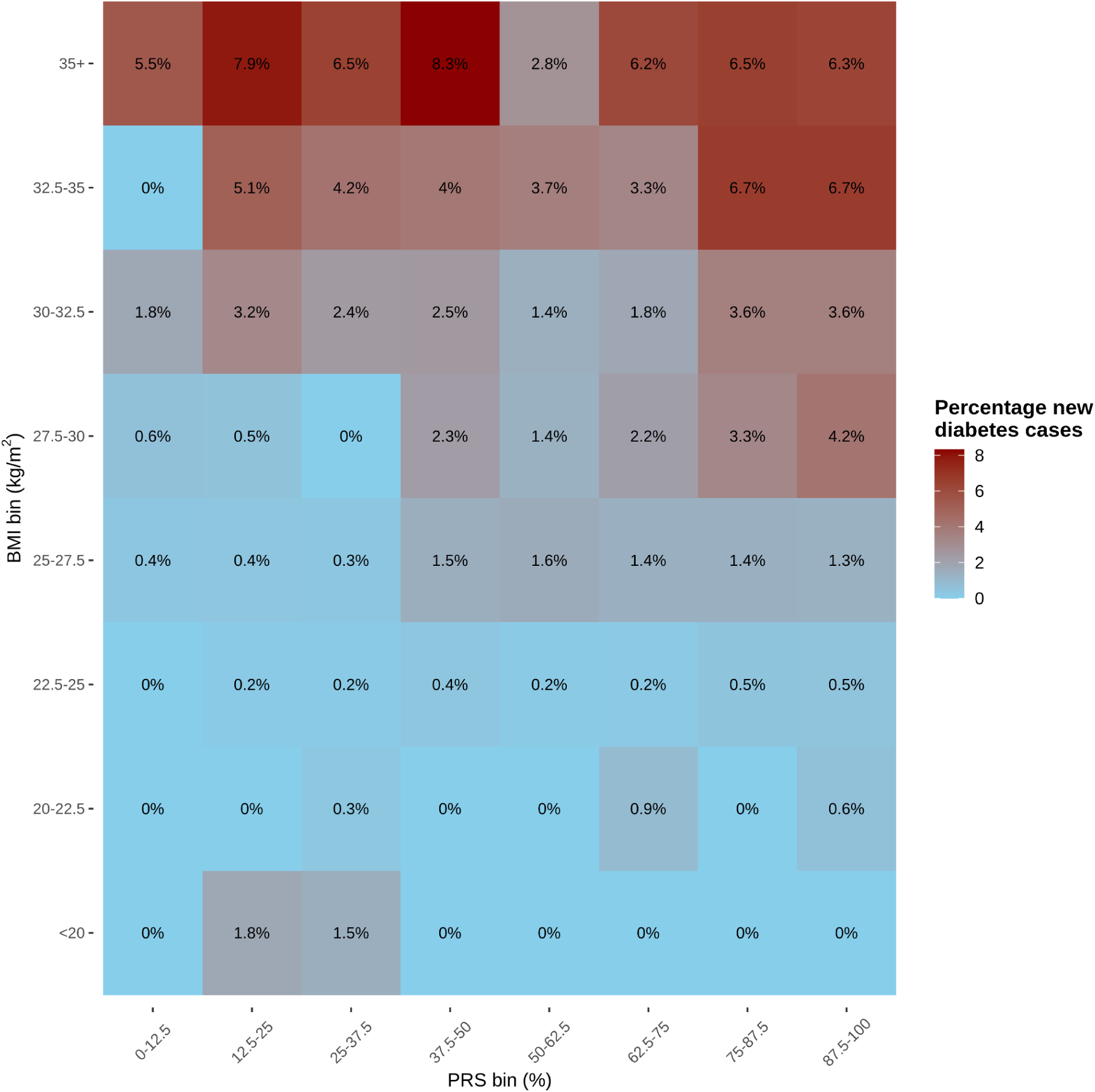
Frequency of developing incident diabetes at four years of follow-up by BMI and T2D PRS. The bins are left-inclusive - e.g. BMI 20-22.5 includes all individuals with a BMI of 20kg/m^2^ to less than 22.5kg/m^2^. T2D PRS derived from 23andMe.

### Sensitivity analyses

Stratifying the analysis by age and sex identified a similar relationship of the PRS for undiagnosed diabetes and prediabetes (**Figures S6A, S6B**) across strata.

While the absolute numbers of undiagnosed and diagnosed diabetes cases increased by strata of the T2D PRS (**Figure S1A**), the ratio of undiagnosed to diagnosed diabetes did not appreciably differ (**Figure S1B**). In contrast, there was a weak trend to an increase in undiagnosed diabetes as a ratio to diagnosed diabetes with increasing BMI (**Figure S2**).

## Discussion

In this study, we demonstrate the utility of a PRS to identify individuals with undiagnosed disease. Compared to conventionally-used measurements of BMI, a T2D PRS identified broadly similar numbers of individuals with undiagnosed diabetes. Importantly, the effects of the T2D PRS and measured weight were additive: when used in addition to individuals with obesity or who are overweight, the combination of measured weight and T2D PRS identified almost all (99%) of individuals with undiagnosed diabetes. This study provides quantitative evidence of the potential clinical utility of PRS in identifying individuals with subclinical disease owing to elevated polygenic risk, providing a new opportunity for implementing genomically-guided precision medicine.

Until recently, diabetes was considered a chronic disease with associated lifelong ailments, including a doubling of risk of mortality and substantial disease-related morbidity^2,21^. This dogma has been recently upended by observational^22–24^ and interventional^25–27^ evidence showing that diabetes can be put into sustained remission^26^ through weight loss. Importantly, prior analyses demonstrate that the sooner interventions geared towards putting diabetes into remission are instituted, the more likely that remission will be achieved^28^. The corollary is that early diagnosis initiatives can identify diabetes that may be more amenable to achieving disease remission. The implication is that individuals with asymptomatic diabetes who achieve remission would never experience the symptoms or consequences of diabetes.

This study provides nascent empirical evidence on the potential utility of PRS at identifying undiagnosed disease in individuals who are not served by existing guidelines^12,13^. T2D is a condition where selecting individuals on the basis of being overweight and having obesity captures the majority (>90%) of individuals with undiagnosed disease in the UK biobank dataset. While use of a PRS, based on a similar number of evaluated individuals as overweight/obesity, increases this value to almost 99%, it is arguable that this is a relatively modest increase in undiagnosed disease identification. However, at the population level, even such modest increases can be of substantial benefit. Furthermore, for any health condition for which a highly informative and easy to measure risk factor such as BMI is unavailable, PRS for that disease may have more unique utility. In the case of diabetes, for those individuals who are at elevated polygenic risk and who have a BMI less than 25 kg/m^2^, use of a PRS has potential clinical value at identifying individuals with undiagnosed disease. For example, current recommendations to screen individuals with obesity or who are overweight^12^ might consider inclusion of a T2D PRS to help further identify individuals with undiagnosed diabetes (as shown in **Table S2**). Current recommendations of age-based screening^11^ might be modified to measure HbA1c at a more frequent cadence and/or starting from an earlier age, among individuals at high phenotypic and genetic risk, in order to detect undiagnosed disease as it emerges.

Our study has several limitations. First, for the purposes of our analysis, which used a PRS evaluated from a European genetic ancestry population in the 23andMe Research Cohort, we restricted the analysis among UKB participants to those individuals who self-reported white ethnicity. It is important that future studies explore the utility of PRS for early diagnosis in non-Europeans, in an effort to bring about equitable precision medicine^29^. Of note, diabetes occurs at lower values of BMI among East Asian individuals as compared to Europeans^30^, meaning it is plausible that the comparative effect of the T2D PRS may be greater in East Asian populations. Second, the recruitment age of the UKB means that we were unable to evaluate the performance of the PRS in individuals younger than 40 years. Adults younger than 35 years represent an age group where PRS may be particularly insightful as it falls outside of the age group (35y+) currently recommended for diabetes screening by ADA^11^ and USPSTF^12^. When stratifying by age, our findings showed a lower absolute number of undiagnosed diabetes cases in younger individuals in the UKB, consistent with the lifecourse epidemiology of diabetes^31^. Nonetheless, previous findings demonstrate a stronger comparative effect of PRS in younger (vs older) individuals, highlighting a clinical space where PRS may be of particular utility^32^. Third, HbA1c screening is not routinely performed in the UK^13^, unlike in the USA, thus the detection rates we identify are unlikely to be directly portable to countries where contemporaneous screening practices, and indeed where the epidemiology of diabetes, differs. Fourth, the self-report in UKB didn’t differentiate between type 1 and type 2 diabetes. However, type 1 diabetes constitutes a small minority (5-10%) of diabetes cases in adulthood^31^, making this an unlikely major source of error in our study. Fifth, it is possible that reverse causality owing to undiagnosed disease led to a reduction in weight, which could inflate the frequency estimates of undiagnosed disease among individuals with low/normal BMI - however a similar pattern was identified for prediabetes where such reverse causation is less likely to manifest. Finally, our PRS explains only a fraction of the heritability, and we’d expect our findings to strengthen as PRS models continue to improve.

In conclusion, we provide quantitative evidence of the application of PRS to early detection of disease. Future studies should expand this to other health conditions, in other contemporary datasets and with a focus on non-European populations. In the era of precision medicine, and with increasing numbers of people seeking direct-to-consumer genotyping, our findings provide empirical evidence in support of interpreting an individual’s genetics to potentially guide initiatives related to early disease detection and thus prevent disease-related morbidity and mortality.

## Data Availability

To be updated at the time of publication.

## Acknowledgements

We thank participants from the UK Biobank and research participants and employees of 23andMe for making this work possible. Chris German, James Ashenhurst, Wei Wang, 23andMe Research Team, Julie M. Granka, Bertram L. Koelsch, Noura S. Abul-Husn, Stella Aslibekyan, Joyce Tung, Suyash S. Shringarpure and Michael V. Holmes are employed by and hold stock or stock options in 23andMe, Inc.

The following members of the 23andMe Research Team contributed to this study:

Stella Aslibekyan, Adam Auton, Elizabeth Babalola, Robert K. Bell, Jessica Bielenberg, Jonathan Bowes, Katarzyna Bryc, Ninad S. Chaudhary, Daniella Coker, Sayantan Das, Emily DelloRusso, Sarah L. Elson, Nicholas Eriksson, Teresa Filshtein, Pierre Fontanillas, Will Freyman, Zach Fuller, Chris German, Julie M. Granka, Karl Heilbron, Alejandro Hernandez, Barry Hicks, David A. Hinds, Ethan M. Jewett, Yunxuan Jiang, Katelyn Kukar, Alan Kwong, Yanyu Liang, Keng-Han Lin, Bianca A. Llamas, Matthew H. McIntyre, Steven J. Micheletti, Meghan E. Moreno, Priyanka Nandakumar, Dominique T. Nguyen, Jared O’Connell, Aaron A. Petrakovitz, G. David Poznik, Alexandra Reynoso, Shubham Saini, Morgan Schumacher, Leah Selcer, Anjali J. Shastri, Janie F. Shelton, Jingchunzi Shi, Suyash Shringarpure, Qiaojuan Jane Su, Susana A. Tat, Vinh Tran, Joyce Y. Tung, Xin Wang, Wei Wang, Catherine H. Weldon, Peter Wilton, Corinna D. Wong.

All authors are employed by and hold stock or stock options in 23andMe, Inc.

This research has been conducted using the UK Biobank Resource under application number 95801.

## References

1. Global Health Estimates: Life expectancy and leading causes of death and disability. The Global Health Observatory. World Health Organization https://www.who.int/data/gho/data/themes/mortality-and-global-health-estimates Accessed Apr 12 2023.

2. Bragg F, Holmes MV, Iona A, et al. Association Between Diabetes and Cause-Specific Mortality in Rural and Urban Areas of China. Jama. 2017;317(3):280–289. doi:10.1001/jama.2016.19720

3. Baena-Díez JM, Peñafiel J, Subirana I, et al. Risk of Cause-Specific Death in Individuals With Diabetes: A Competing Risks Analysis. Diabetes Care. 2016;39(11):1987–1995. doi:10.2337/dc16-0614

4. Diabetes Key Facts. World Health Organization https://www.who.int/news-room/fact-sheets/detail/diabetes Accessed Apr 12 2023.

5. Collaborators G 2021 D, Ong KL, Stafford LK, et al. Global, regional, and national burden of diabetes from 1990 to 2021, with projections of prevalence to 2050: a systematic analysis for the Global Burden of Disease Study 2021. Lancet. 2023;402(10397):203-234. doi:10.1016/s0140-6736(23)01301-6

6. National Diabetes Statistics Report: Prevalence of Both Diagnosed and Undiagnosed Diabetes. National Diabetes Statistics Report Centers for Disease Control and Prevention website https://www.cdc.gov/diabetes/data/statistics-report/index.html Accessed Apr 6 2023. https://www.cdc.gov/diabetes/data/statistics-report/diagnosed-undiagnosed-diabetes.html

7. Fang M, Wang D, Coresh J, Selvin E. Undiagnosed Diabetes in U.S. Adults: Prevalence and Trends. Diabetes Care. 2022;45(9):1994–2002. doi:10.2337/dc22-0242

8. Laiteerapong N, Ham SA, Gao Y, et al. The Legacy Effect in Type 2 Diabetes: Impact of Early Glycemic Control on Future Complications (The Diabetes & Aging Study). Diabetes Care. 2019;42(3):416–426. doi:10.2337/dc17-1144

9. Kim KJ, Choi J, Bae JH, et al. Time to Reach Target Glycosylated Hemoglobin Is Associated with Long-Term Durable Glycemic Control and Risk of Diabetic Complications in Patients with Newly Diagnosed Type 2 Diabetes Mellitus: A 6-Year Observational Study. Korean Diabetes J. 2020;45(3):368–378. doi:10.4093/dmj.2020.0046

10. Khunti K, Aroda VR. Coming Full Circle: Prioritizing Early Glycemic Control to Reduce Microvascular and Macrovascular Complications in People With Type 2 Diabetes. Diabetes Care. 2022;45(4):766–768. doi:10.2337/dci21-0064

11. ElSayed NA, Aleppo G, Aroda VR, et al. 2. Classification and Diagnosis of Diabetes: Standards of Care in Diabetes—2023. Diabetes Care. 2022;46(Supplement_1):S19-S40. doi:10.2337/dc23-s002

12. Force UPST, Davidson KW, Barry MJ, et al. Screening for Prediabetes and Type 2 Diabetes. Jama. 2021;326(8):736–743. doi:10.1001/jama.2021.12531

13. Adult Screening Programme Diabetes. UK National Screening Committee Screening Recomendation https://view-health-screening-recommendations.service.gov.uk/diabetes/.

14. Ashenhurst JR, Sazonova OV, Svrchek O, et al. A Polygenic Score for Type 2 Diabetes Improves Risk Stratification Beyond Current Clinical Screening Factors in an Ancestrally Diverse Sample. Front Genet. 2022;13:871260. doi:10.3389/fgene.2022.871260

15. Bycroft C, Freeman C, Petkova D, et al. The UK Biobank resource with deep phenotyping and genomic data. Nature. 2018;562(7726):203-209. doi:10.1038/s41586-018-0579-z

16. Tung JY, Do CB, Hinds DA, et al. Efficient Replication of over 180 Genetic Associations with Self-Reported Medical Data. Plos One. 2011;6(8):e23473. doi:10.1371/journal.pone.0023473

17. Ashenhurst JR*, Zhan J*, Multhaup ML, et al. White Paper 23-21 A Generalized Method for the Creation and Evaluation of Polygenic Scores. 23andMe, Inc https://permalinks23andme.com/pdf/23_21-PRSMethodology_May2020.pdf. Published online October 1, 2020. https://permalinks.23andme.com/pdf/23_21-PRSMethodology_May2020.pdf

18. Cropano C, Santoro N, Groop L, et al. The rs7903146 Variant in the TCF7L2 Gene Increases the Risk of Prediabetes/Type 2 Diabetes in Obese Adolescents by Impairing β-Cell Function and Hepatic Insulin Sensitivity. Diabetes Care. 2017;40(8):1082–1089. doi:10.2337/dc17-0290

19. Pedregosa F, Varoquaux G, Gramfort A, et al. Scikit-learn: Machine Learning in Python. Journal of Machine Learning Research. 2011;12:2925–2830.

20. Arch BN, Blair J, McKay A, Gregory JW, Newland P, Gamble C. Measurement of HbA1c in multicentre diabetes trials – should blood samples be tested locally or sent to a central laboratory: an agreement analysis. Trials. 2016;17(1):517. doi:10.1186/s13063-016-1640-6

21. Yang JJ, Yu D, Wen W, et al. Association of Diabetes With All-Cause and Cause-Specific Mortality in Asia. Jama Netw Open. 2019;2(4):e192696. doi:10.1001/jamanetworkopen.2019.2696

22. Gregg EW, Chen H, Wagenknecht LE, et al. Association of an Intensive Lifestyle Intervention With Remission of Type 2 Diabetes. Jama. 2012;308(23):2489–2496. doi:10.1001/jama.2012.67929

23. Buchwald H, Avidor Y, Braunwald E, et al. Bariatric Surgery: A Systematic Review and Meta-analysis. Jama. 2004;292(14):1724–1737. doi:10.1001/jama.292.14.1724

24. Arterburn DE, Bogart A, Sherwood NE, et al. A Multisite Study of Long-term Remission and Relapse of Type 2 Diabetes Mellitus Following Gastric Bypass. Obes Surg. 2013;23(1):93–102. doi:10.1007/s11695-012-0802-1

25. Lean ME, Leslie WS, Barnes AC, et al. Primary care-led weight management for remission of type 2 diabetes (DiRECT): an open-label, cluster-randomised trial. Lancet. 2018;391(10120):541–551. doi:10.1016/s0140-6736(17)33102-1

26. Lean MEJ, Leslie WS, Barnes AC, et al. Durability of a primary care-led weight-management intervention for remission of type 2 diabetes: 2-year results of the DiRECT open-label, cluster-randomised trial. Lancet Diabetes Endocrinol. 2019;7(5):344–355. doi:10.1016/s2213-8587(19)30068-3

27. Geltrude M, Simona P, Andrea DG, et al. Bariatric Surgery versus Conventional Medical Therapy for Type 2 Diabetes. New Engl J Med. 2012;366(17):1577–1585. doi:10.1056/nejmoa1200111

28. Karter AJ, Nundy S, Parker MM, Moffet HH, Huang ES. Incidence of Remission in Adults With Type 2 Diabetes: The Diabetes & Aging Study. Diabetes Care. 2014;37(12):3188–3195. doi:10.2337/dc14-0874

29. Martin AR, Kanai M, Kamatani Y, Okada Y, Neale BM, Daly MJ. Clinical use of current polygenic risk scores may exacerbate health disparities. Nat Genet. 2019;51(4):584–591. doi:10.1038/s41588-019-0379-x

30. Ma RCW, Chan JCN. Type 2 diabetes in East Asians: similarities and differences with populations in Europe and the United States. Ann Ny Acad Sci. 2013;1281(1):64–91. doi:10.1111/nyas.12098

31. Bullard KM, Cowie CC, Lessem SE, et al. Prevalence of Diagnosed Diabetes in Adults by Diabetes Type — United States, 2016. Morbidity Mortal Wkly Rep. 2018;67(12):359-361. doi:10.15585/mmwr.mm6712a2

32. Jiang X, Holmes C, McVean G. The impact of age on genetic risk for common diseases. Plos Genet. 2021;17(8):e1009723. doi:10.1371/journal.pgen.1009723

33. Lee JJ, Wedow R, Okbay A, et al. Gene discovery and polygenic prediction from a genome-wide association study of educational attainment in 1.1 million individuals. Nat Genet. 2018;50(8):1112–1121. doi:10.1038/s41588-018-0147-3

