## Supplemental material for "A polygenic risk score identifies undiagnosed cases of diabetes"

Author Affiliations:

### Supplementary Figures and Tables

**Figure S1. Undiagnosed and diagnosed diabetes by strata of T2D PRS: (A) absolute counts; (B) as a proportion to total diabetes cases**

A:

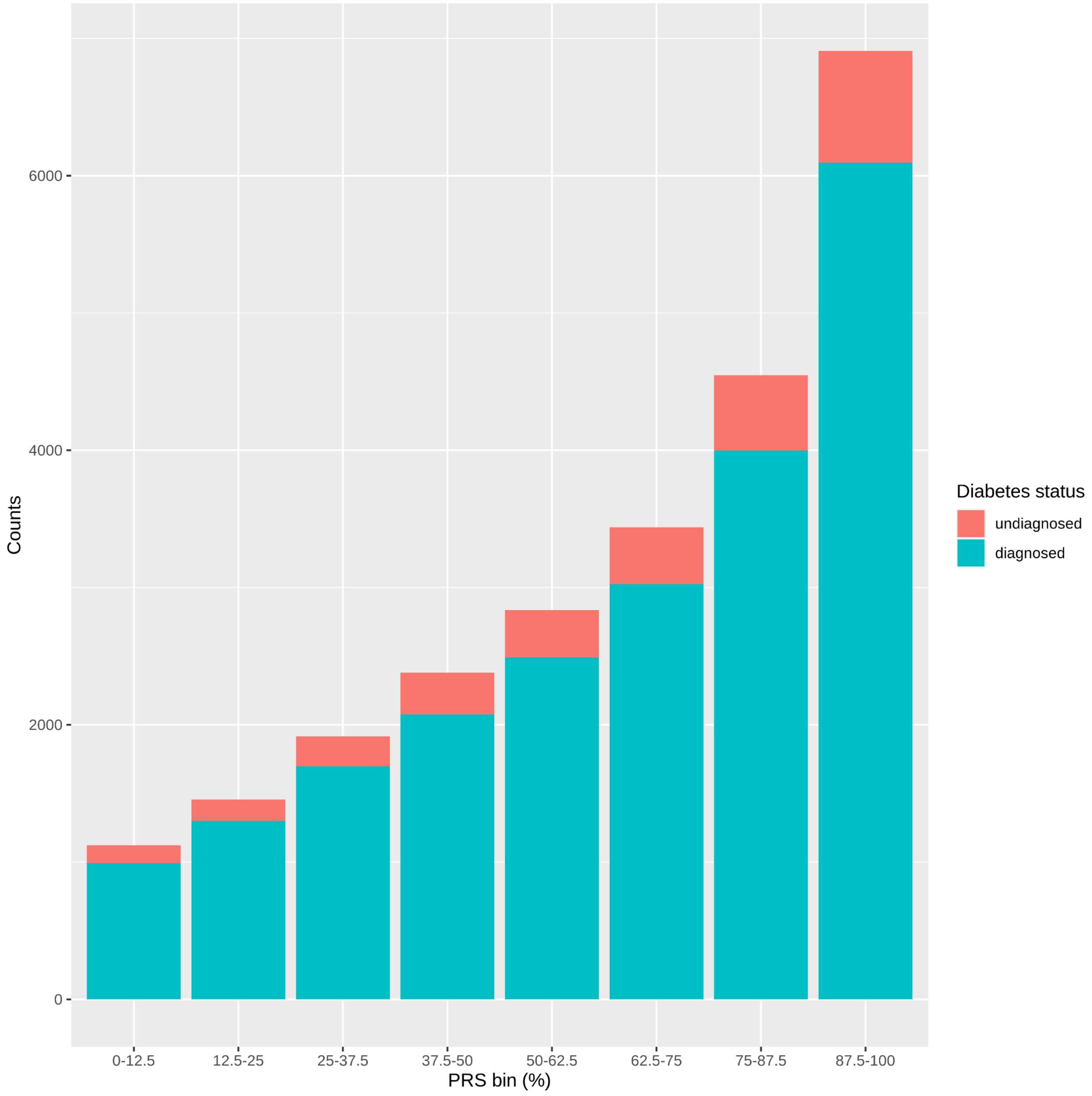

B:

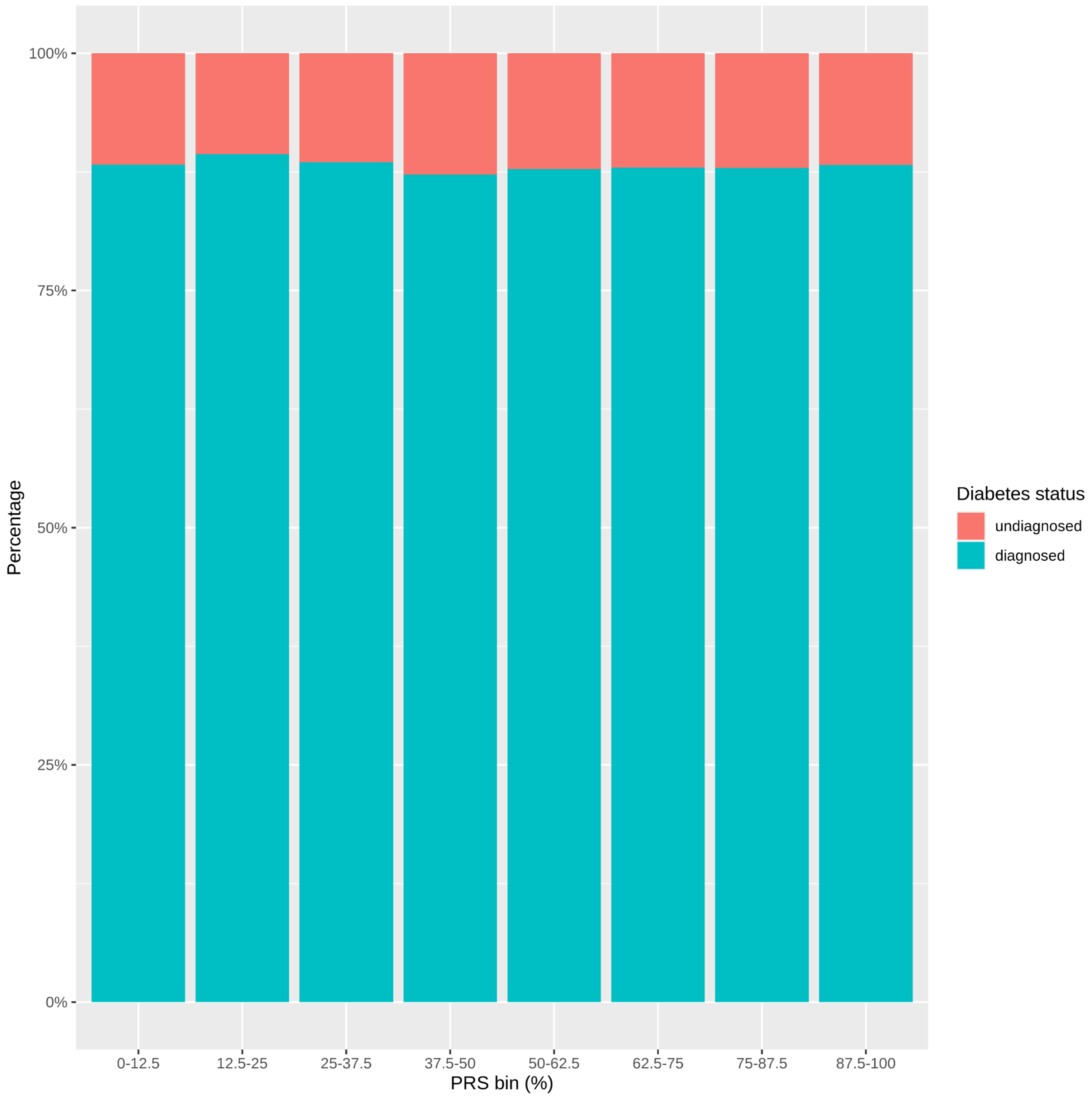

100%
75%
50%
25%
0%
Percentage
0-12.5
12.5-25
25-37.5
37.5-50
50-62.5
62.5-75
75-87.5
87.5-100
PRS bin (%)
Diabetes status
undiagnosed
diagnosed

**Figure S2.Undiagnosed and diagnosed diabetes by strata of measured BMI: (A) absolute counts; (B) as a proportion to total diabetes cases**

A:

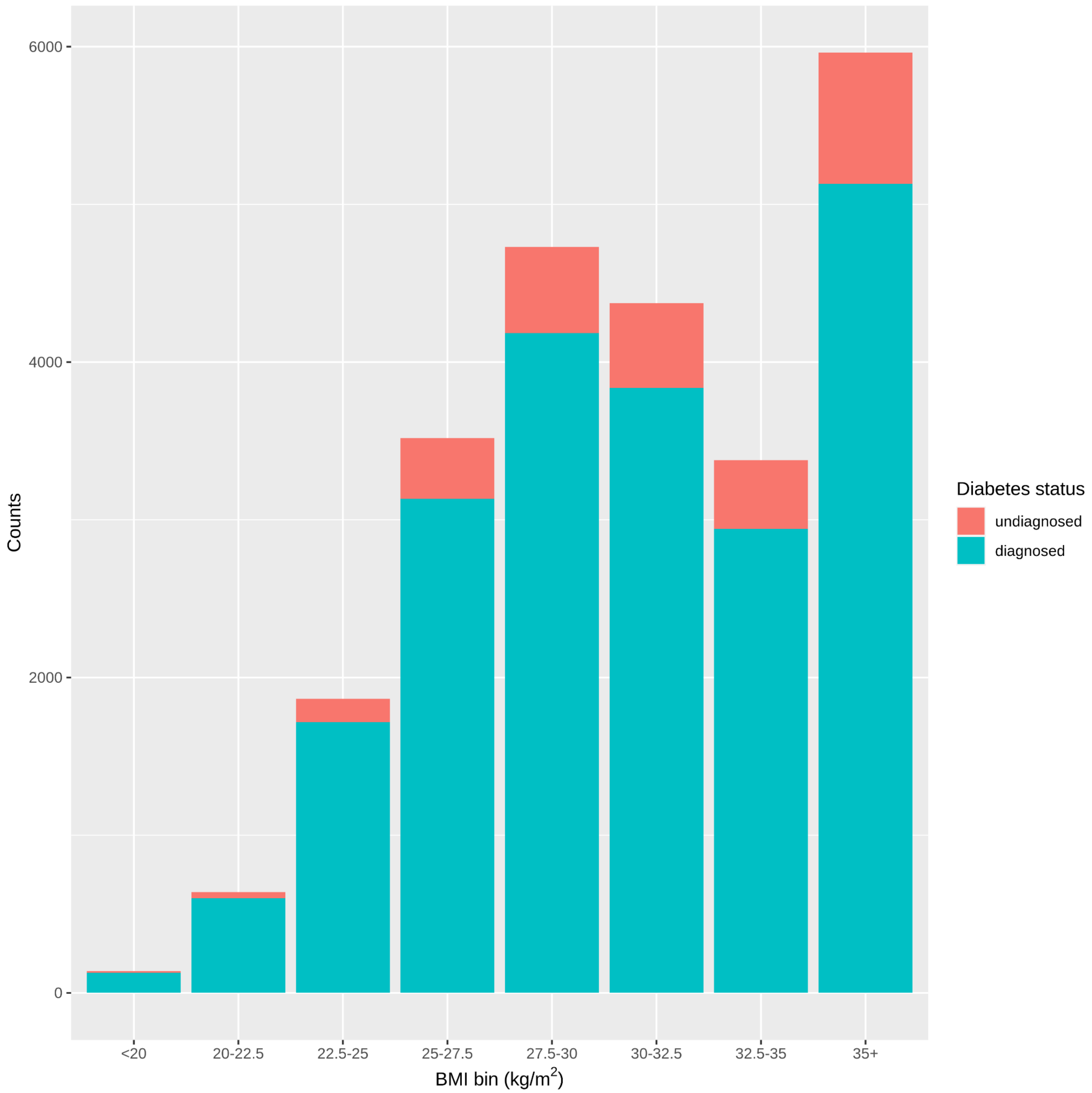

B:

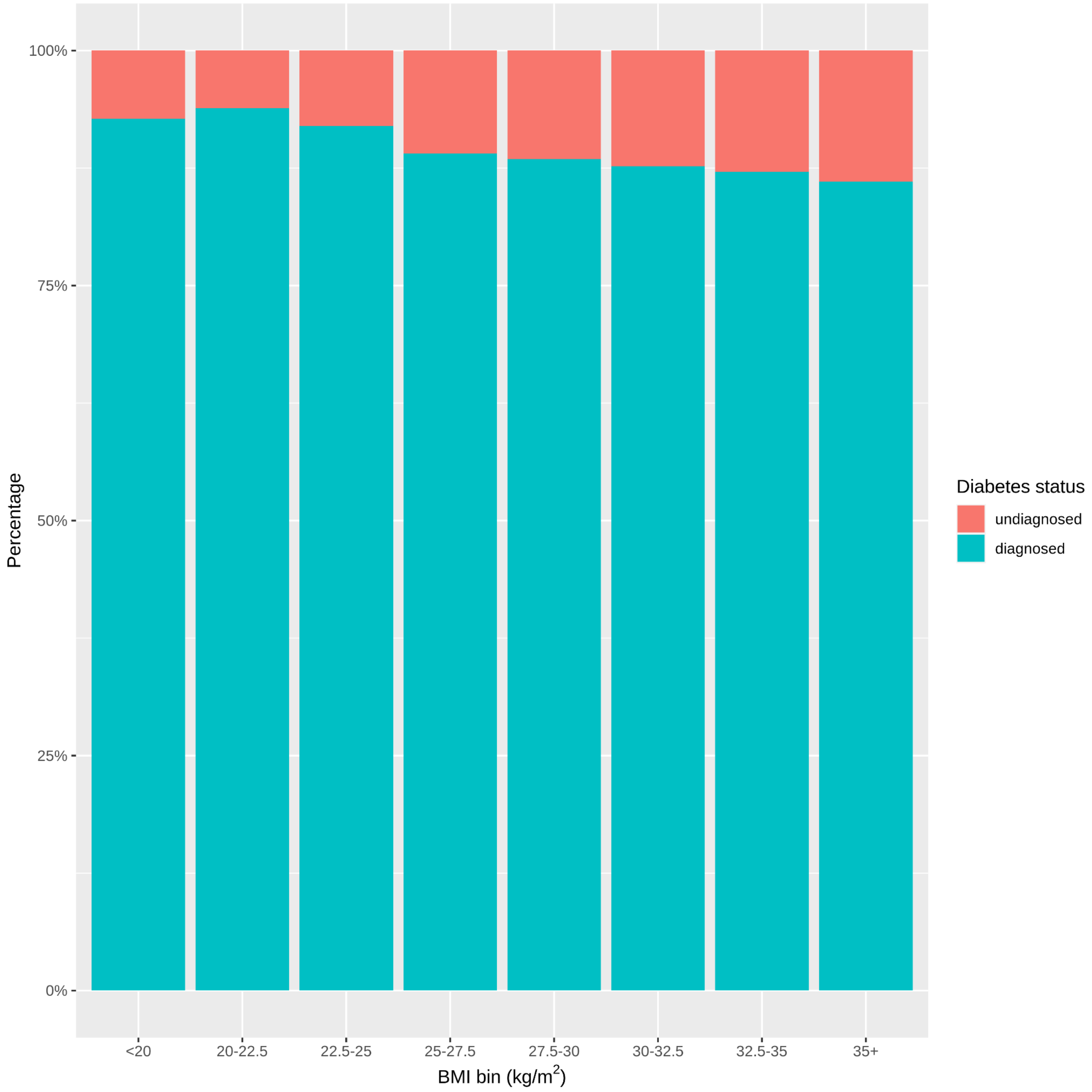

100%
75%
50%
25%
0%
Percentage
<20
20-22.5
22.5-25
25-27.5
27.5-30
30-32.5
32.5-35
35+
BMI bin (kg/m2)
Diabetes status
undiagnosed
diagnosed

**Figure S3. 2D bar chart plot of undiagnosed diabetes by T2D PRS and BMI. Error bars relate to 95% confidence intervals.**

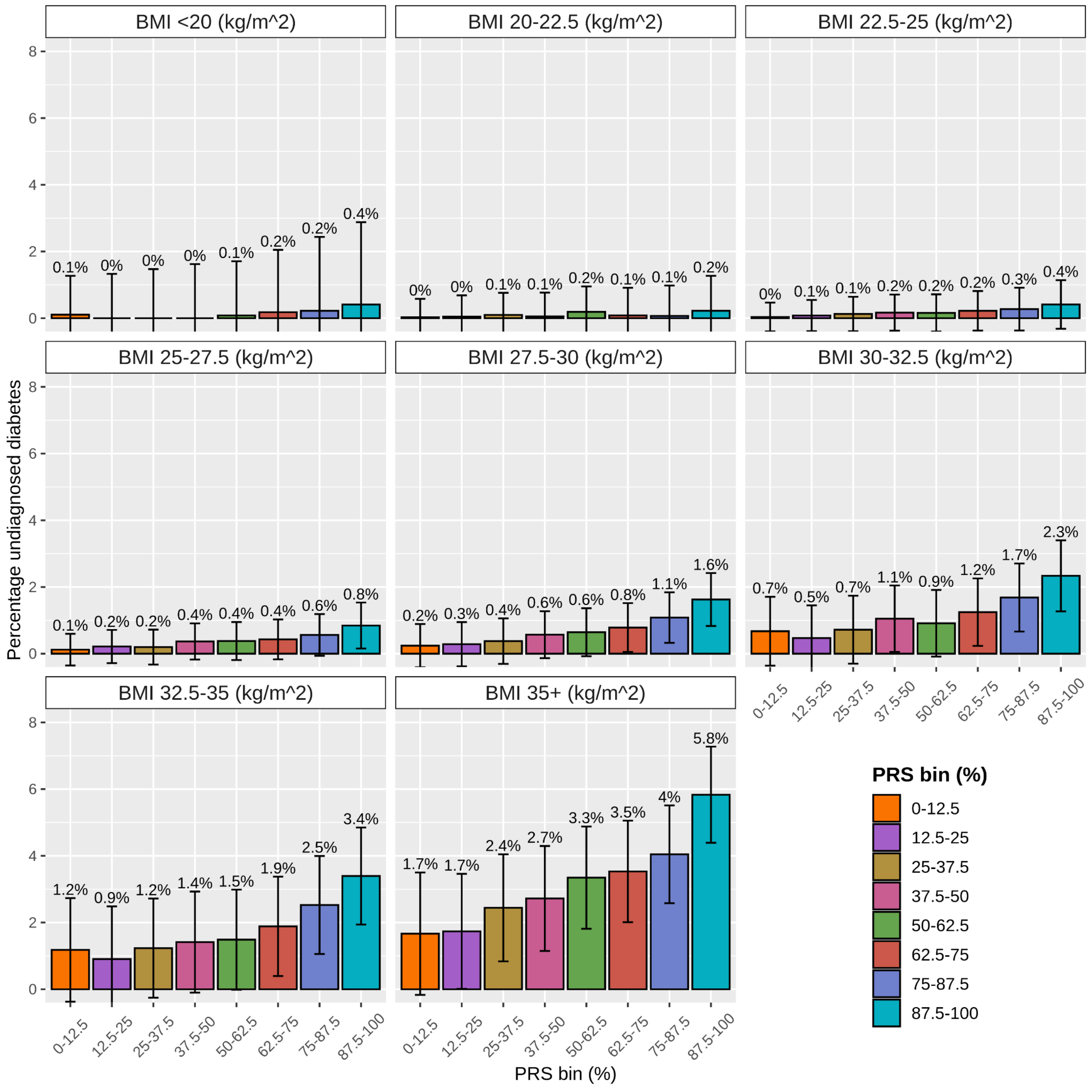

**Figure S4. Proportion of undiagnosed diabetes cases detected by a T2D PRS and BMI, displayed by a heatmap plot.**

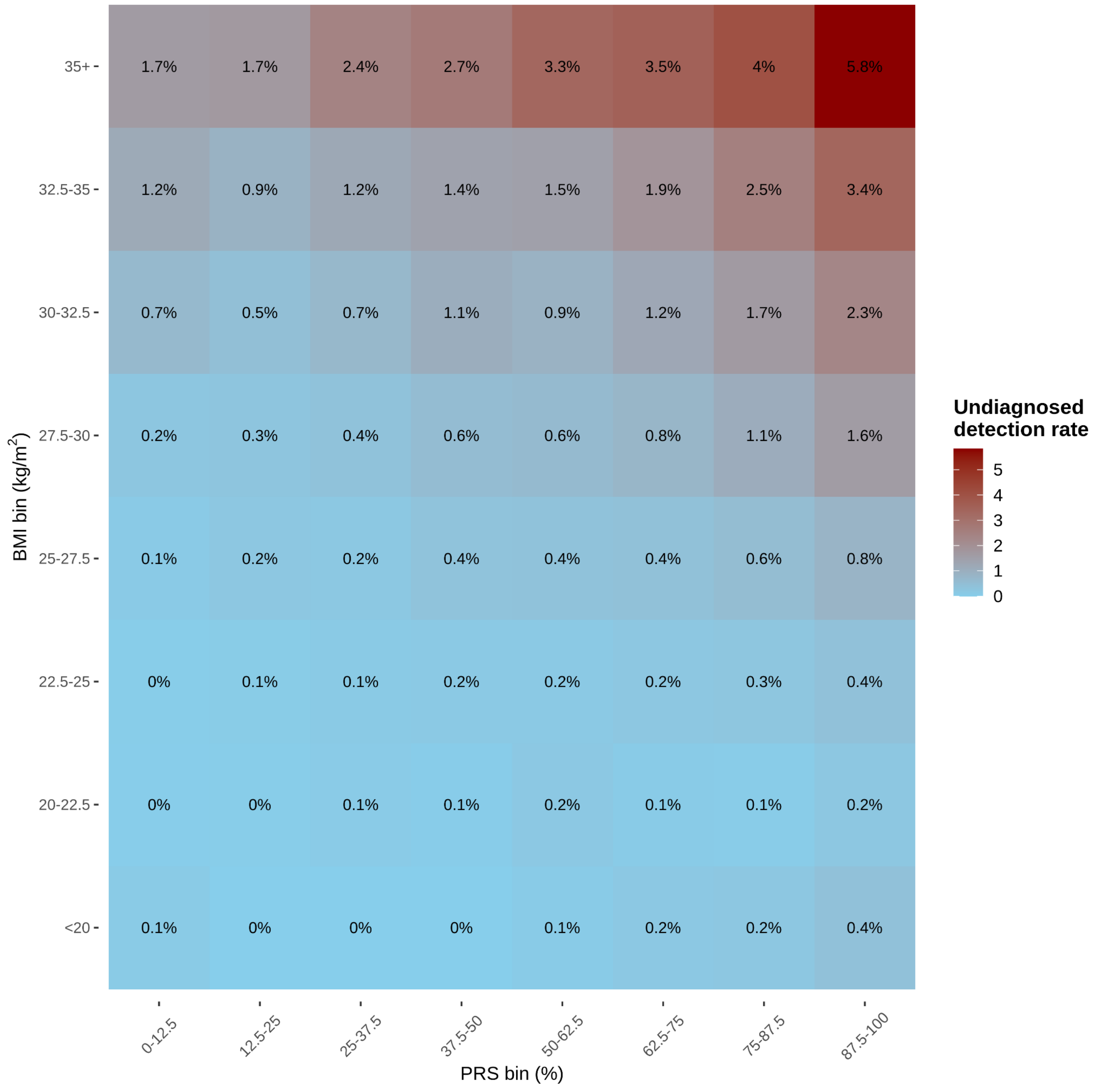

**Figure S5. Proportion of prediabetes cases detected by a T2D PRS and BMI, displayed by a heatmap plot.**

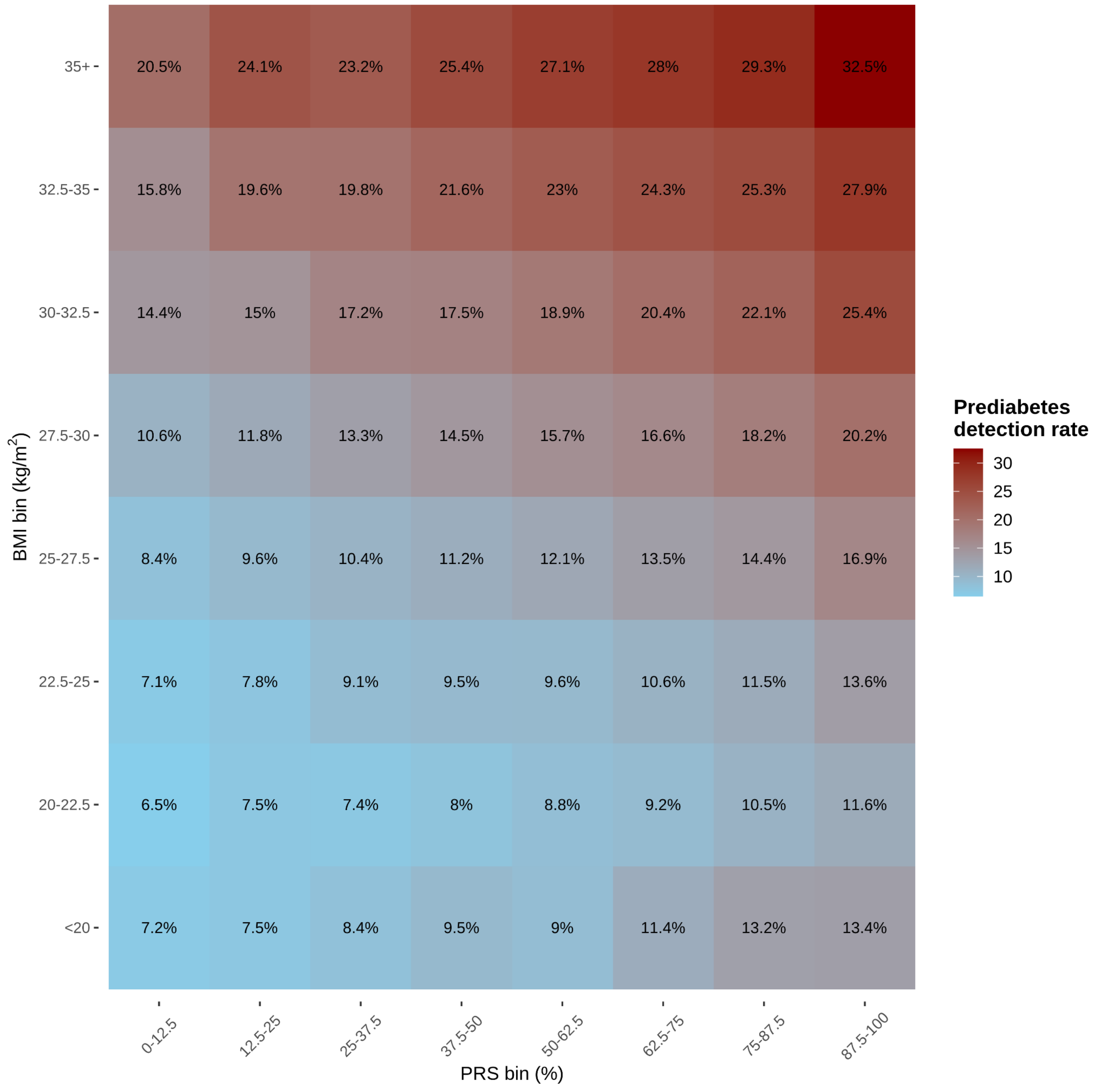

**Figure S6**. **Probability of undiagnosed diabetes by strata of T2D PRS, stratified by (A) sex and (B) age.**

A:

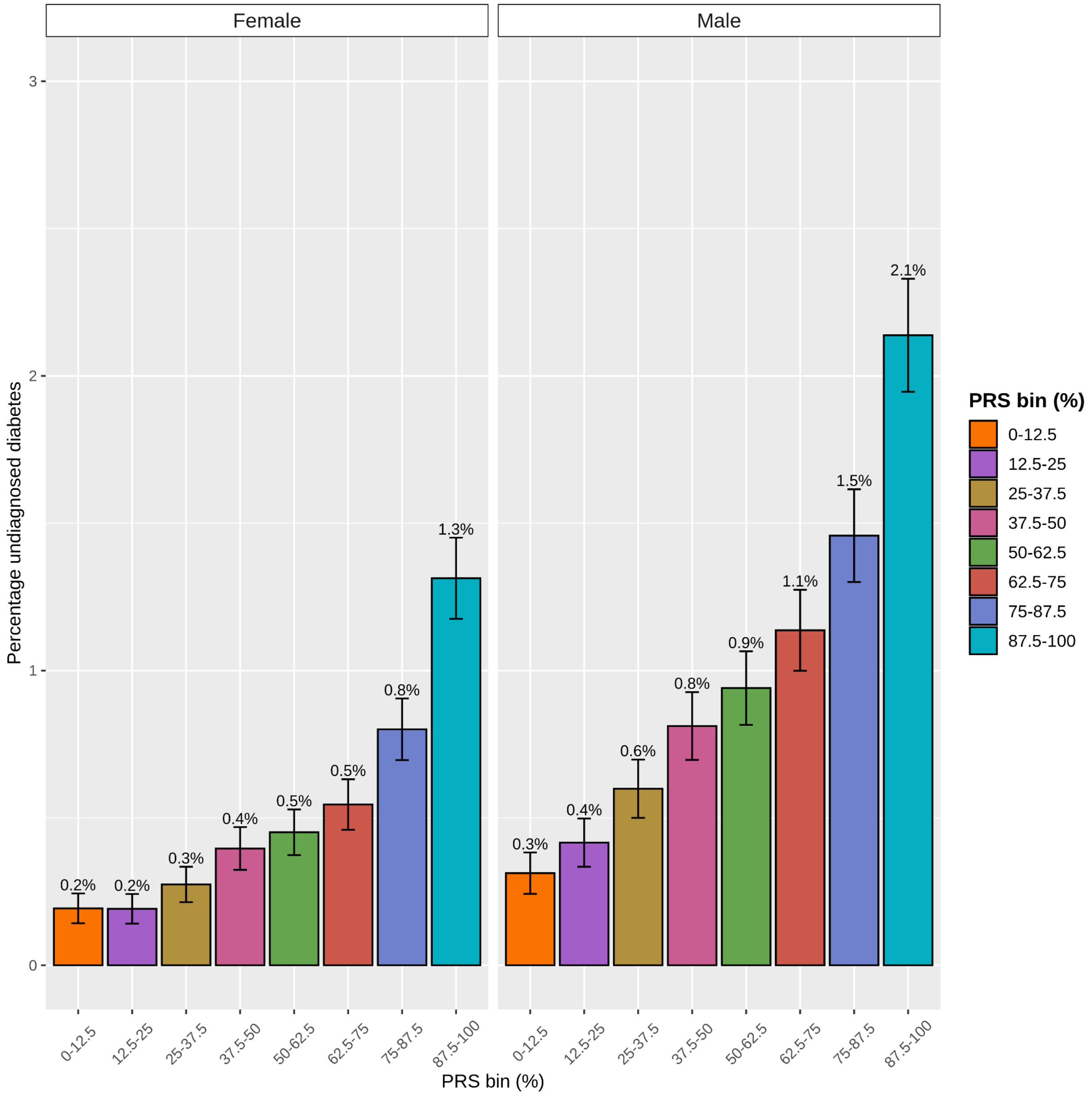


B:

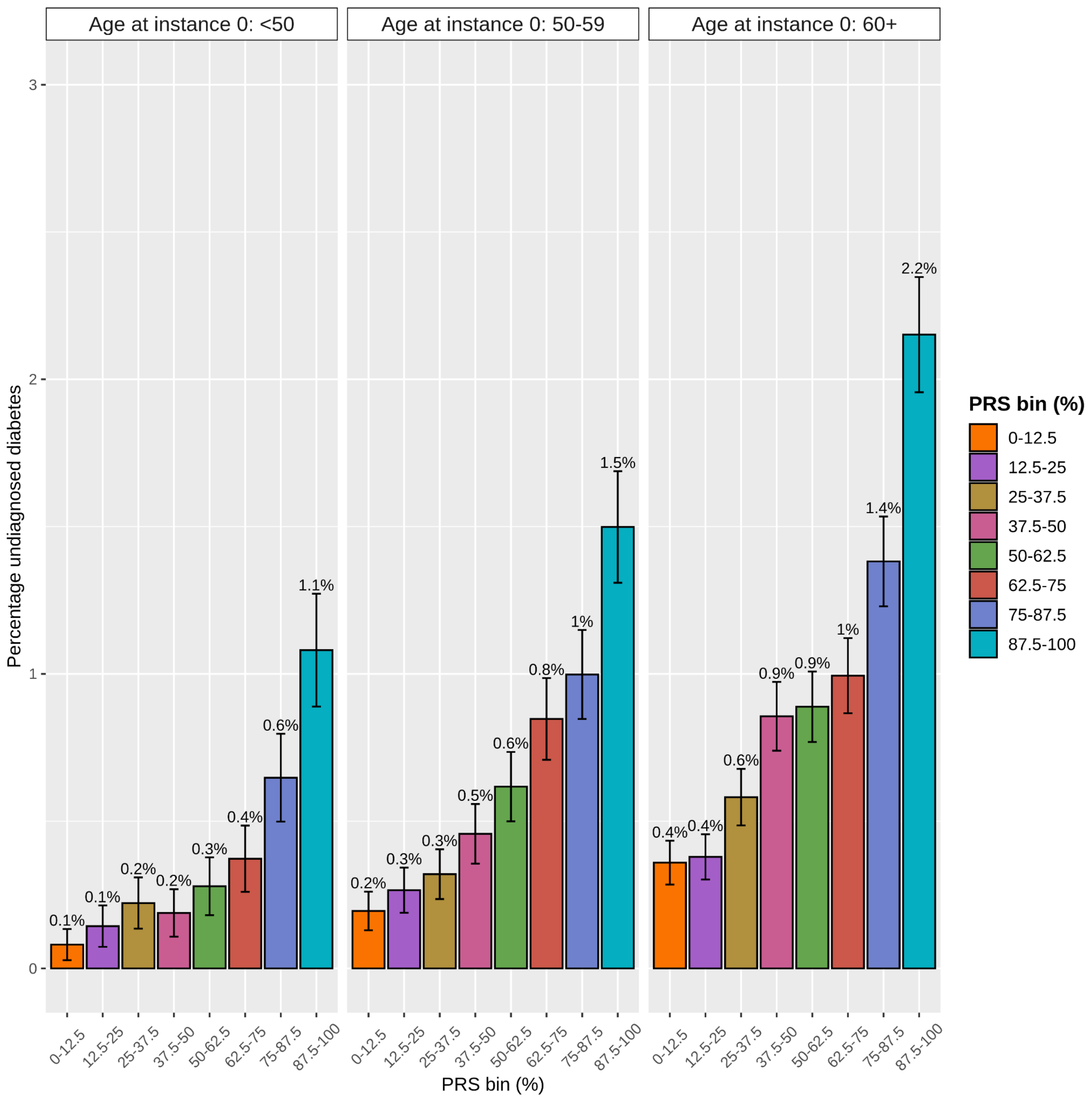

Age at instance 0: <50
Age at instance 0: 50-59
Age at instance 0: 60+
Percentage undiagnosed diabetes
3
2
1
0
0.1%
0.1%
0.2%
0.2%
0.3%
0.4%
0.6%
1.1%
0.2%
0.3%
0.3%
0.5%
0.6%
0.8%
1%
1.5%
0.4%
0.4%
0.6%
0.9%
0.9%
1%
1.4%
2.2%
0-12.5
12.5-25
25-37.5
37.5-50
50-62.5
62.5-75
75-87.5
87.5-100
PRS bin (%)
PRS bin (%)
0-12.5
12.5-25
25-37.5
37.5-50
50-62.5
62.5-75
75-87.5
87.5-100

**Figure S7**. **Density plot of T2D PRS values in the UK Biobank by diabetes status.**

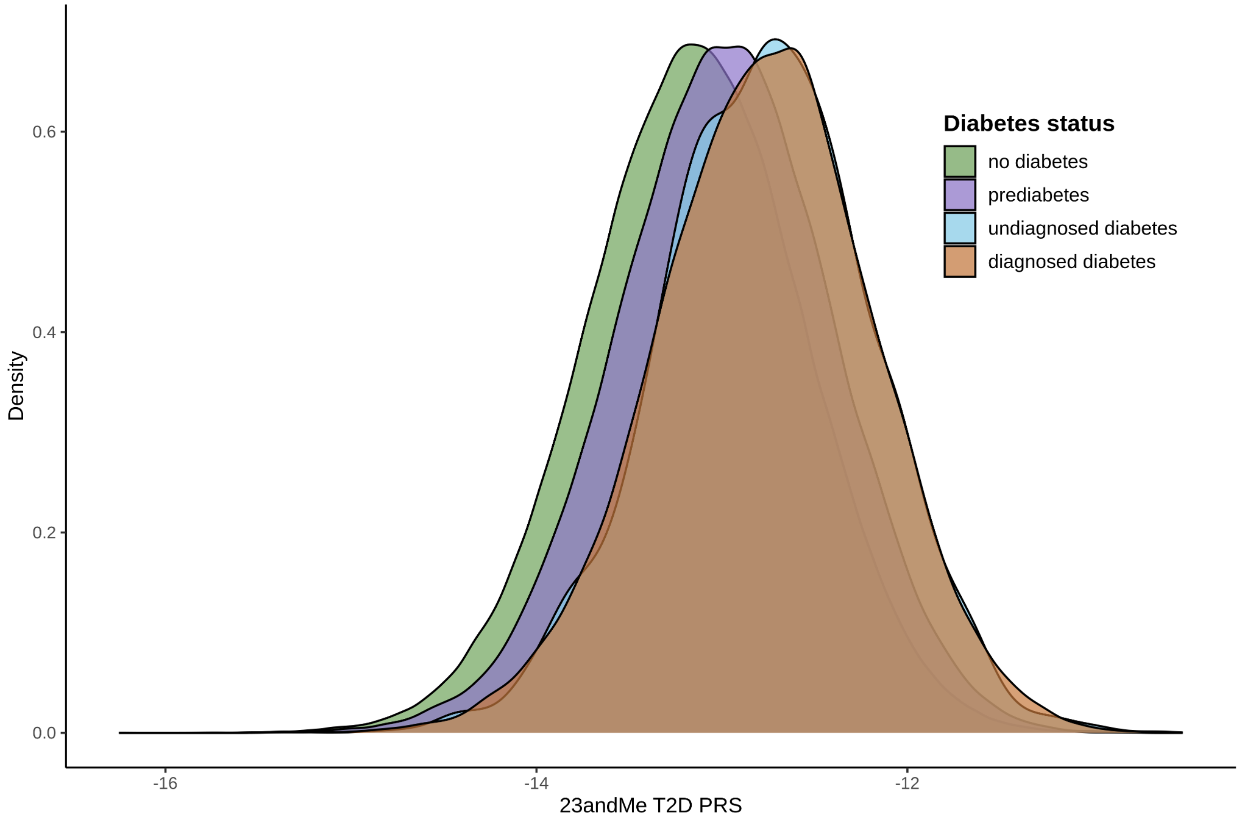

**Figure S8**. **Plot of the empirical cumulative distribution function (eCDF) of HbA1c among individuals without doctor-diagnosed diabetes at study baseline.** The blue dashed line indicates the threshold for prediabetes levels and the red dashed line indicates the threshold for diabetes.

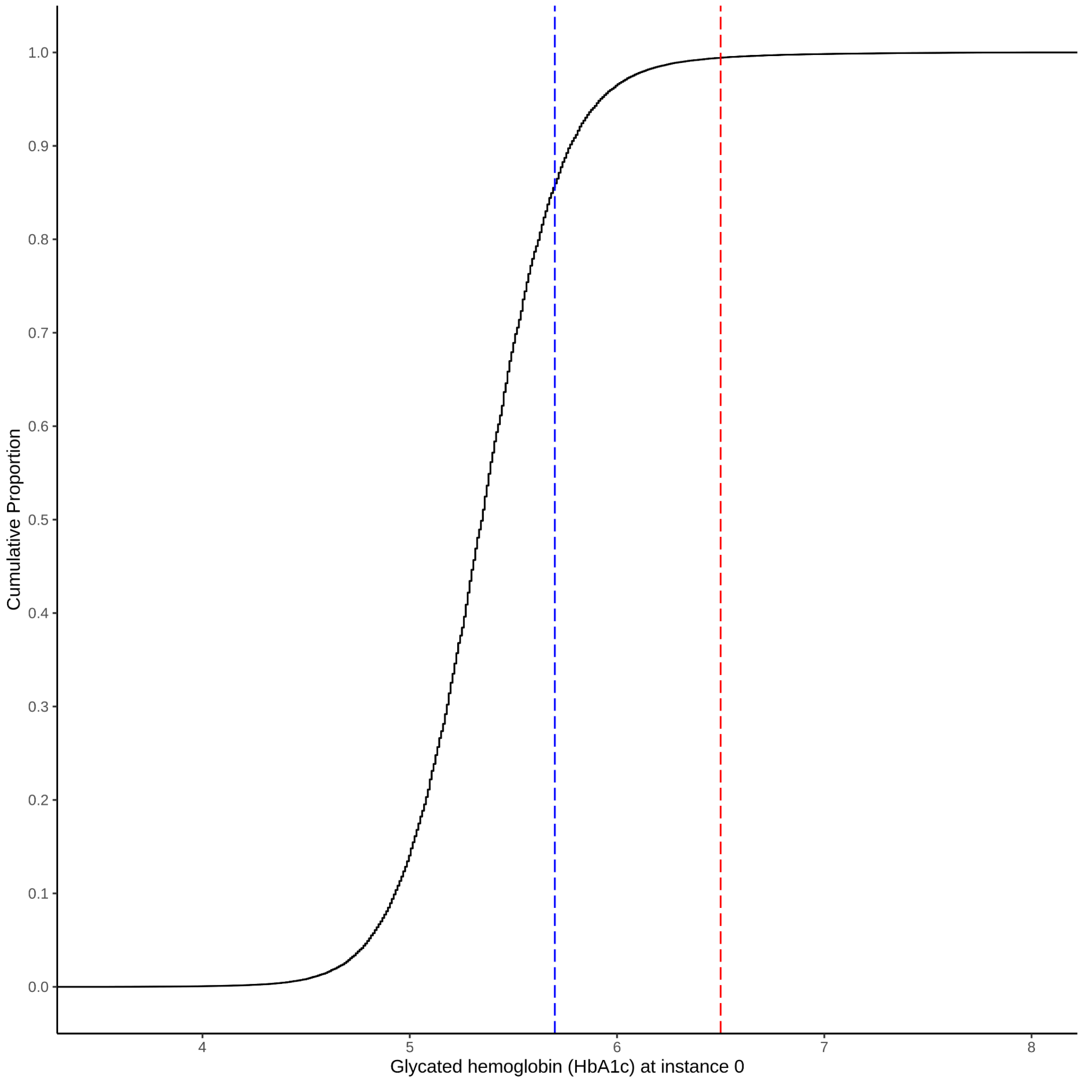

**Table S1. Data field definitions.** Definitions of data fields used from the first occurrence data (UK Biobank Category 1712).

| Data field ID | Data field concept |
|---|---|
| 130706 | Date E10 first reported (insulin-dependent diabetes mellitus) |
| 130707 | Source of report of E10 (insulin-dependent diabetes mellitus) |
| 130708 | Date E11 first reported (non-insulin-dependent diabetes mellitus) |
| 130709 | Source of report of E11 (non-insulin-dependent diabetes mellitus) |
| 130710 | Date E12 first reported (malnutrition-related diabetes mellitus) |
| 130711 | Source of report of E12 (malnutrition-related diabetes mellitus) |
| 130712 | Date E13 first reported (other specified diabetes mellitus) |
| 130713 | Source of report of E13 (other specified diabetes mellitus) |
| 130714 | Date E14 first reported (unspecified diabetes mellitus) |
| 130715 | Source of report of E14 (unspecified diabetes mellitus) |

Footnote: E10-E14 refer to ICD-10 codes (defined in the brackets)

**Table S2. Additive effect of T2D PRS in addition to overweight/obesity to identify individuals with undiagnosed diabetes.** Analyses were restricted to individuals who self-report an absence of doctor diagnosed diabetes and who have an HbA1c measurement, both at baseline.

| BMI | T2D PRS | Undiagnosed diabetes (% total undiagnosed) |
|---|---|---|
| ≥25 $kg/m^2$ | - | 93% |
| ≥25 $kg/m^2$ | Top 5% | 94% |
| ≥25 $kg/m^2$ | Top 14% | 95% |
| ≥25 $kg/m^2$ | Top 26% | 96% |
| ≥25 $kg/m^2$ | Top 41% | 97% |
| ≥25 $kg/m^2$ | Top 54% | 98% |
| ≥25 $kg/m^2$ | Top 69% | 99% |

**Table S3. Comparison of measured BMI and a T2D PRS at detecting undiagnosed diabetes when normalized to the same number of individuals that have a BMI ≥25$kg/m^2$.** Analyses were restricted to individuals who self-report an absence of doctor diagnosed diabetes and who have an HbA1c measurement, both at baseline

| | BMI ≥25$kg/m^2$ (BMI in the top 65.6%) | T2D PRS in the top 65.6% |
|---|---|---|
| Number with HbA1c ≥ 6.5% at instance 0 | 2,735 | 2,504 |
| Number with HbA1c < 6.5% at instance 0 | 267,758 | 267,989 |
| Detection rate (%) | 1.01 | 0.93% |
| Proportion (%) of undiagnosed diabetes detected | 93.2% | 85.3% |
